# Pre-procedural testing improves estimated COVID-19 prevalence and trends

**DOI:** 10.1101/2022.04.13.22273200

**Authors:** Genevieve C. Pang, Amy T. Hou, Krizhna L. Bayudan, Ethan A. Frank, Jennifer Pastiglione, Lorrin W. Pang

## Abstract

**Background:** COVID-19 positivity rates reported to the public may provide a distorted view of community trends because they tend to be inflated by high-risk groups, such as symptomatic patients and individuals with known exposures to COVID-19. This positive bias within high-risk groups has also varied over time, depending on testing capability and indications for being tested. In contrast, throughout the pandemic, routine COVID-19 screening tests for elective procedures and operations unrelated to COVID-19 risk have been administered by medical facilities to reduce transmission to medical staffing and other patients. We propose the use of these pre-procedural COVID-19 patient datasets to reduce biases in community trends and better understand local prevalence.

**Methods:** Using patient data from the Maui Medical Group clinic, we analyzed 12,640 COVID-19 test results from May 1, 2020 to March 16, 2021, divided into two time periods corresponding with Maui’s outbreak.

**Results:** Mean positivity rates were 0.1% for the pre-procedural group, 3.9% for the symptomatic group, 4.2% for the exposed group, and 2.0% for the total study population. Post-outbreak, the mean positivity rate of the pre-procedural group was significantly lower than the aggregate group (all other clinic groups combined). The positivity rates of both pre-procedural and aggregate groups increased over the study period, although the pre-procedural group showed a smaller rise in rate.

**Conclusions:** Pre-procedural groups may produce different trends compared to high-risk groups and are sufficiently robust to detect small changes in positivity rates. Considered in conjunction with high-risk groups, pre-procedural marker groups used to monitor understudied, low-risk subsets of a community may improve our understanding of community COVID-19 prevalence and trends.

## 1. Background

Reported COVID-19 positivity rates likely provide a distorted view of community trends because they tend to be inflated by high-risk groups. During the COVID-19 pandemic, when diagnostic testing capacity was scarce, symptomatic and exposed patients were preferentially tested to describe and control outbreaks^1^. Other high priority groups included travelers and individuals in contact with clusters of cases (e.g., local prisons and elderly facilities). This approach to testing likely contributed to a positive bias in COVID-19 positivity rates. These rates were also prone to temporal inconsistencies in testing capacity, algorithms, and methods (e.g., rapid antigen versus polymerase-chain reaction). For example, as voluntary diagnostic testing became freely available to the public, rates were prone to a self-selection bias, as positive results led to enforced isolation. Prevalence estimates are used to make inferences about latent community prevalence and are critical drivers of public perception of COVID-19 and public health policies^2^. Left unaddressed, biases within these estimates may result in poor-fitting public-health policies and legislations for COVID-19.

What methods could improve the accuracy of community COVID-19 prevalence and epidemic trends? Ideally, to determine the latent prevalence of COVID-19 in a community, a large group of individuals would be sampled randomly and tested irrespective of factors positively or negatively associated with COVID-19. However, random sampling is logistically difficult under the constraints of a pandemic, and such studies are often affected by low participation rates and self-selection bias ^3-6^. Prior studies have investigated alternative approaches to reduce bias in estimated prevalence, using mathematical models with randomized pooling^4-5,7^ and online incentivized experiments to estimate prevalence based on key COVID-19 risk factors^3^.

We propose that biases within COVID-19 positivity rates may be reduced empirically by recalculating rates from existing datasets. Across the country, medical facilities require COVID-19 screening tests before elective procedures or operations to prevent transmission to medical staffing and other patients. These datasets for pre-operative (i.e., pre-procedural) patient groups may offer key insights regarding COVID-19 rates and trends because they tend to be 1) robust, especially for large hospitals and clinics, 2) already collected and readily available for analysis, and 3) prone to fewer testing indications with respect to COVID-19 risk when compared to high-risk groups.

## 2. Data and Methods

The Maui District Health Office division under the Hawaii Department of Health partnered with the Maui Medical Group (i.e., MMG), a multi-site, multi-specialty clinic with a 35,000-40,000-patient base on Maui (approximately 20-25% of Maui’s population). The MMG provided 12,630 COVID-19 test results using de-identified electronic medical records for all patients tested from May 1, 2020 - March 16, 2021. Patients were tested by the SARS-CoV-2 polymerase-chain reaction nucleic acid amplification test. This dataset included the age, gender, city of residence, facility, primary care provider, reason for testing, and date of test, at two-week intervals.

Based on the chief reason for testing, patients were sorted into seven mutually exclusive groups: pre-procedural, symptomatic, exposed, travel-related, unspecified, occupational, and healthcare-related (Figure 1). In the rare instance that a patient listed multiple reasons for testing in addition to symptoms, the patient was sorted into the symptomatic group. Positivity rates (i.e., number of positive cases divided by number of total tests) were calculated for individual groups and the study population (i.e., all groups combined) at two-week intervals and over the total study period.

**Figure 1.**
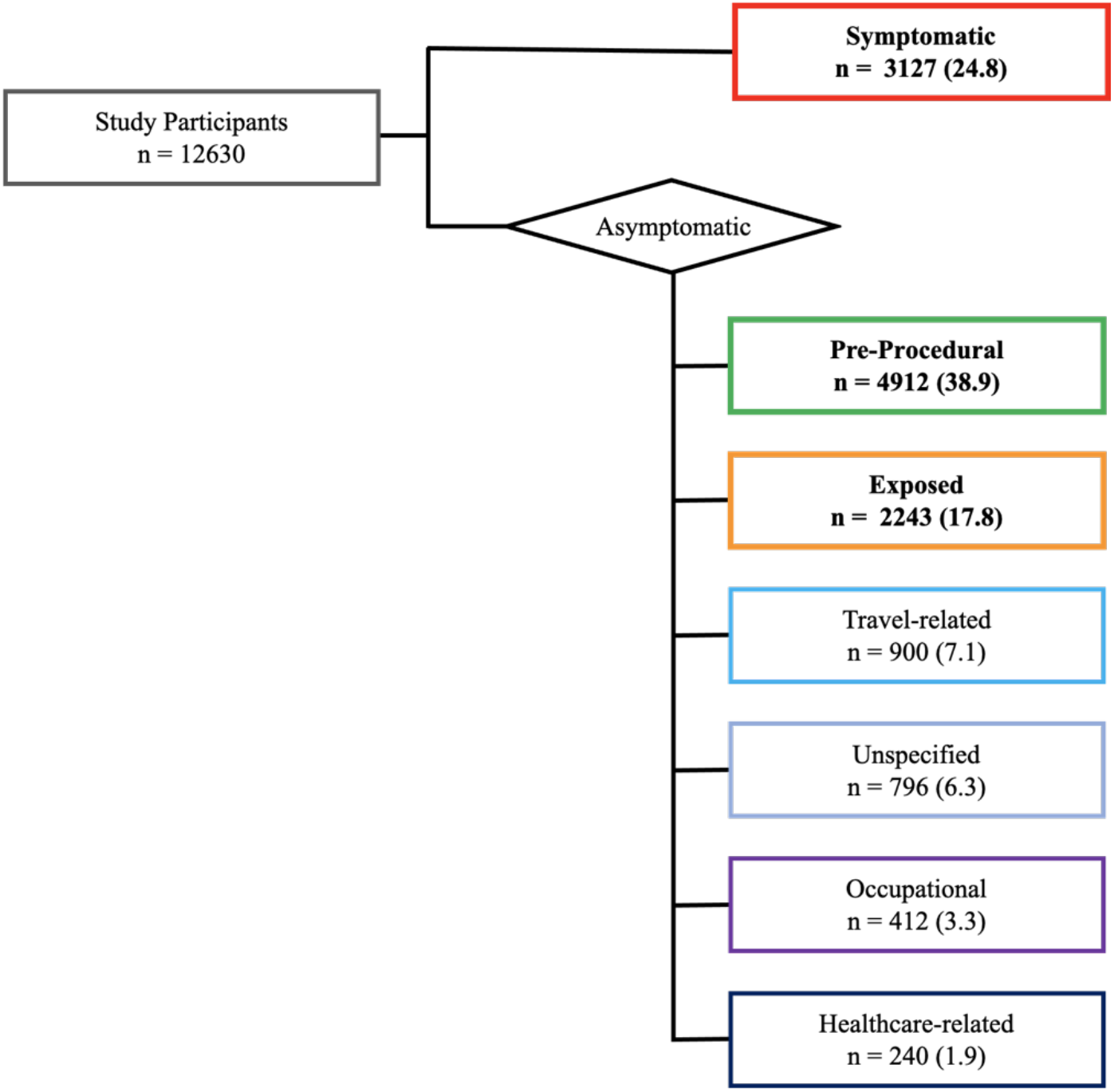
Flow diagram of study participants. Participants tested for COVID-19 are categorized into seven groups: symptomatic (red), pre-procedural (green), exposed (orange), travel-related (light blue), unspecified (medium blue), occupational (purple), and healthcare-related (dark blue). No. (%).

The **pre-procedural group** included individuals who required clearance for elective procedures in gastroenterology (e.g., colonoscopies), orthopedic surgery (e.g., hip and knee replacements), general surgery (e.g.., cholecystectomies, lipoma removals, and lacerations), gynecology, and otolaryngology (e.g.., bronchoscopies). Appointments were scheduled approximately 10-25 days in advance. Pre-procedural screening tests were performed within 72 hours of the scheduled procedure. Patients who tested negative were cleared for procedures. Patients who tested positive were placed in isolation, with results sent to the Maui District Health Office, and their procedures were either canceled or rescheduled. For the island of Maui, elective procedures were paused at the beginning of the pandemic and resumed on April 25, 2020.

The **symptomatic group** included individuals with one or more symptoms associated with COVID-19 (e.g., fever, cough, sore throat, shortness of breath, nausea, vomiting, abdominal pain, loss of smell or loss of taste). The **exposed group** included individuals exposed to suspected COVID-positive cases. The **travel-related group** included individuals tested for travel-related reasons (e.g., pre-departure, ongoing travel, and post-arrival testing). The **unspecified group** included individuals who requested a test for no specified reason. The **occupational group** included individuals who required a test for non-healthcare establishments (e.g., K-12 schools, colleges, sports teams, child and adult daycares, and group homes). The **healthcare-related group** included individuals associated with a healthcare facility (e.g., staff at outpatient clinics or hospitals, patients at rehabilitation, dialysis, or food-disorder treatment centers, and companions to those undergoing medical procedures).

COVID-19 testing volumes and positivity rates were calculated at two-week intervals for the study population and its individual groups. We compared positivity rates among larger groups (i.e., pre-procedural, symptomatic, and exposed), where n > 2000, to determine how individual groups may contribute to the trend in the MMG population. We assessed whether the pre-procedural group differed from other groups to determine the effects of bias on positivity rates and trends among groups over time. We compared the demographics of this study’s population and its groups to the demographics of the general MMG clinic population, by age ranges and genders (Table 1). We also compared the positivity rates and trends of MMG population to those of Maui County and Hawaii state.

**Table 1.** Demographic characteristics of study groups (pre-procedural, symptomatic, exposed, travel, request, occupational risk, and healthcare setting), combined MMG study groups (study population), and all patients (tested and untested) served by MMG (clinic population) sampled from 5/1/2020-3/16/2021.

| Characteristic | Pre-Procedural<br>N = 4827 | Symptomatic<br>N = 3065 | Possible Exposure<br>N = 2179 | Travel<br>N = 873 | Request<br>N = 794 | Occupational Risk<br>N = 401 | Healthcare Setting<br>N = 231 | Study Population<br>N = 12370 | Clinic Population<br>N = 45132 |
| --- | --- | --- | --- | --- | --- | --- | --- | --- | --- |
| Gender - no. (%) |  |  |  |  |  |  |  |  |  |
| Male | 2453 (50.8) | 1299 (42.4) | 989 (45.4) | 397 (45.5) | 355 (44.7) | 195 (48.6) | 88 (38.1) | 5776 (46.7) | 20812 (46.1) |
| Female | 2374 (49.2) | 1766 (57.6) | 1190 (54.6) | 476 (54.5) | 439 (55.3) | 206 (51.4) | 143 (61.9) | 6594 (53.3) | 2430 (53.9) |
| Age Group - no. (%) |  |  |  |  |  |  |  |  |  |
| 0-17 | 111 (2.3) | 406 (13.2) | 308 (14.1) | 78 (8.9) | 69 (8.7) | 28 (7.0) | 2 (0.9) | 1002 (8.1) | 8614 (19.1) |
| 18-29 | 183 (3.8) | 540 (17.6) | 406 (18.6) | 157 (18.0) | 103 (13.0) | 124 (30.9) | 27 (11.7) | 1540 (12.5) | 4356 (9.7) |
| 30-44 | 524 (10.9) | 721 (23.5) | 518 (23.8) | 190 (21.8) | 180 (22.7) | 112 (27.9) | 87 (37.7) | 2332 (18.9) | 6923 (15.3) |
| 45-59 | 1408 (29.2) | 646 (21.1) | 507 (23.2) | 238 (27.3) | 191 (24.1) | 80 (20.0) | 55 (23.8) | 3125 (25.3) | 9133 (20.2) |
| 60-75 | 2090 (43.3) | 599 (19.5) | 366 (16.8) | 188 (21.5) | 192 (24.2) | 40 (10.0) | 41 (17.7) | 3516 (28.4) | 11775 (26.1) |
| 75+ | 511 (10.6) | 153 (5.0) | 74 (3.4) | 22 (2.5) | 59 (7.4) | 17 (4.2) | 19 (8.2) | 855 (6.9) | 4331 (9.6) |
| Positive Tests - no. (%) |  |  |  |  |  |  |  |  |  |
|  | 6 (0.1) | 122 (3.9) | 95 (4.2) | 6 (0.7) | 12 (1.5) | 11 (2.7) | 1 (0.2) | 253 (2.0) | - |

To statistically compare positivity rates between groups we used Chi-Square tests for homogeneity and Fisher’s exact tests for rare outcomes. We compared positivity rates of pre-procedural and the combined non-pre-procedural groups (i.e., aggregate group) at two-week intervals (Table 3a). To test for trends over time, we compared positivity rates calculated for the first and second study time periods (May 2020 through November 2020 and November 2020 through March 2021, respectively), corresponding with Maui’s outbreak pattern, within and between the pre-procedural and aggregate groups.

The Hawaii Department of Health’s Institutional Review Board reviewed this study as a project in partnership with MMG. The project was approved and qualified as exempt research under 45 CFR 46.116(d) of the Department of Health and Human Services, waiving informed consent based on the minimal risk associated with de-identified data used in this study.

## 3. Results

Positivity rates were estimated using 12,630 COVID-19 test results from the Maui Medical Group clinic, collected from May 2020 - March 2021. Patients reported the following reasons for COVID-19 testing, in descending order: pre-procedural testing (38.9%), symptomatic (24.8%), exposed (17.8%), travel-related (7.1%), unspecified (6.3%), occupational (3.3%), and healthcare-related (1.9%) (Figure 1). The pre-procedural group comprised approximately 40% of the total study population (Figure 2). The average positivity rates of the groups over the study period were 2.0% (MMG study population), 0.1% (pre-procedural), 3.9% (symptomatic), 4.2% (exposed), 0.7% (travel-related), 1.5% (unspecified), 2.7% (occupational), and 0.4% (healthcare-related).

**Figure 2.**
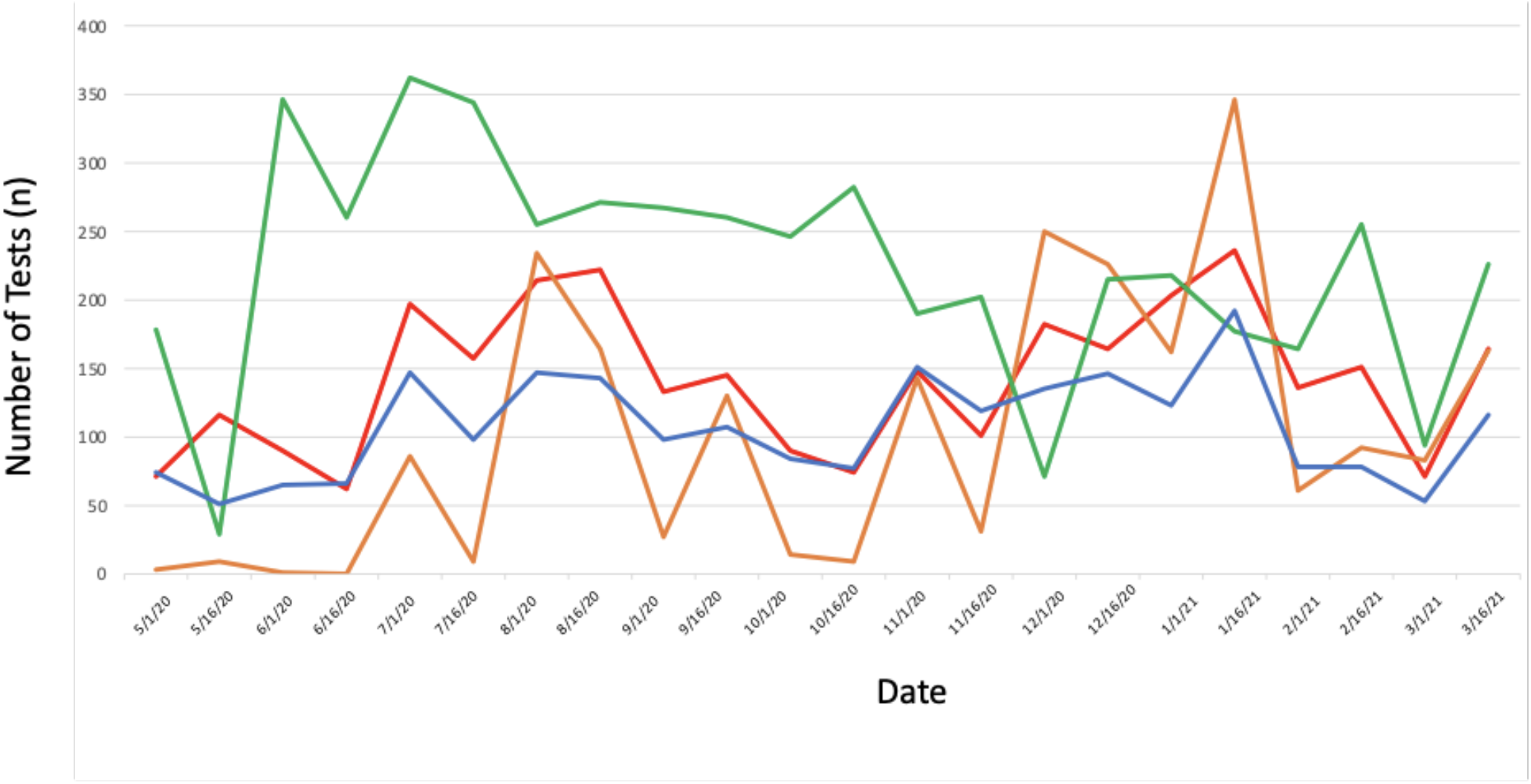
Number of tests at two-week intervals. COVID-19 tests collected by MMG every two weeks for symptomatic (red), exposed (orange), pre-procedural (green) groups, and remaining groups combined (blue).

The positivity rates of the pre-procedural group were significantly lower than the aggregate group, both at specific two-week intervals (Figure 3a) and through the first and the second time periods of this study (Figure 3a, P < 10^−5^ for each period, Fisher’s exact test). Between the first and second time periods, the positivity rate of the aggregate group significantly increased (P < 10^−5^), as did the positivity rate of the pre-procedural group, which showed a small but significant increase (P = .01) from 0% (0/25720) to 0.26% (6/2340).

**Figure 3.**
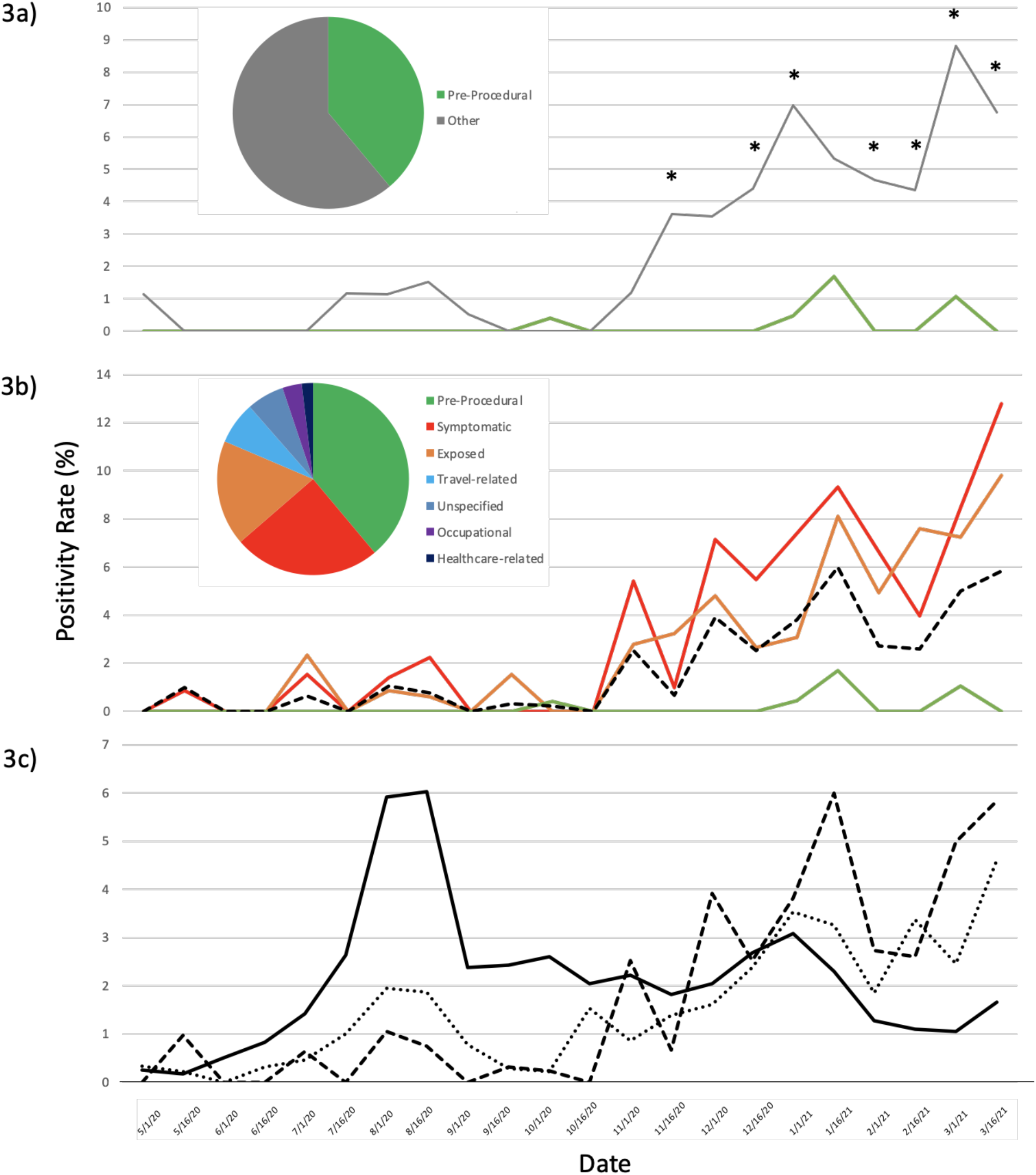
Comparisons of COVID-19 positivity rates at two-week intervals 3a) between the MMG pre-procedural group (green) and remaining MMG groups combined (gray), 3b) among the MMG population (dashed black), symptomatic group (red), exposed group (orange), and pre-procedural group (green), and 3c) among the MMG population (dashed), Maui County (dotted), and Hawaii state (solid). Pie charts separate the MMG population into 3a) pre-procedural and aggregate groups and 3b) individual MMG groups. Statistically significant differences in positivity rate at two-week intervals are indicated with an asterisk (*).

The positivity rates of the MMG population were similar to those of the high-risk, symptomatic and exposed groups (Figure 3b). COVID-19 positivity rates of MMG, Maui County, and Hawaii state showed similar trends between MMG and Maui County, and a roughly inverted pattern at the state level (Figure 3c).

## 4. Discussion

The results of this study show how high-risk groups can drive reported COVID-19 positivity rates. For MMG, positive COVID-19 cases from high-risk groups greatly influenced the positivity rate of its population, resulting in more similar trends among these groups compared to the pre-procedural group. This finding shows how combining data into a single metric to describe the positivity rate of a community, irrespective of sampling strategy, can distort positivity rates. The magnitude of these distortions would also likely vary over time, based on changing testing resources and policies. For example, when resources are scarce, more stringent testing requirements would focus on the most high-risk cases, increasing the positivity rates of tested groups.

We emphasize that, while absolute values of percent positivity from pre-procedural groups are not accurate measures of latent community prevalence, pre-procedural datasets may improve our ability to accurately detect changes in COVID-19 community trends. This study’s analysis of the pre-procedural group was statistically powerful enough to detect a significant rise (0-0.26%) in its positivity rate from the first to second time period, despite much lower positivity rates relative to other groups. This finding indicates that pre-procedural data are sufficiently robust to detect relatively incremental changes in positivity rates over time. Pre-procedural groups may thus be a suitable marker to investigate trends and relative changes in positivity rates, an approach that has been successfully established for other diseases such as HIV^2^.

Over the course of this study, the overall trend in the positivity rates of the pre-procedural and aggregate groups were similar: both indicated a rise in the positivity rate of COVID-19 from the first to second time period. However, the magnitude of the rise was lower for the pre-procedural group than the aggregate group. Additionally, a peak in positivity rate occurred during the first time period for the aggregate group but not the pre-procedural group; this peak likely reflects changes in testing capacity or policies, versus a change in latent community prevalence. The observed differences in COVID-19 trends between this study’s pre-procedural and aggregate groups show how an overrepresentation of high-risk groups within datasets can result in very different patterns of community prevalence. We argue that pre-procedural groups provide key insights about a large, neglected, lower-risk subset of the community, and should be considered when estimating COVID-19 latent community prevalence and trends.

In a similar study conducted in Detroit, Michigan, 10 out of 4,381 (0.4%) pre-procedural patients tested positive for COVID-19^8^. In contrast, during the study period, the city of Detroit reported much higher daily positivity rates (2.9-9.8%)^9^. The authors concluded that, in settings with limited diagnostic resources, testing should be prioritized for symptomatic patients because asymptomatic, low-risk pre-procedural patients had relatively much lower positivity rates compared to high-risk groups. We propose a different interpretation; large discrepancies between the estimated positivity rates of low- and high-risk groups indicates a greater need for the inclusion of low-risk (e.g., pre-procedural) groups to improve the accuracy of estimated COVID-19 community prevalence.

The findings of this study suggest that larger scale trends for the total MMG study population, Maui County, and Hawaii state likely contain distortions caused by changes in testing protocols over time and the disproportionate sampling of high-risk groups. The MMG population trend generally matched that of Maui County, where the positivity rate slowly trended upwards. In contrast, the trend for Hawaii state shows a very different pattern with peaks in July-August 2020 followed by a plateau. Outbreak control may have contributed to the apparent dramatic increases in cases but, if transmission were successfully contained, may misrepresent the epidemiologic trend for Hawaii state. Future studies could expand upon this study to compare statewide pre-procedural patient data with reported positivity rates.

### Study limitations

Although pre-procedural groups do not contain the same biases as high-risk groups, this group may not approximate a randomized sample in several critical ways. First, pre-procedural patients may behave more cautiously and limit potential exposures in anticipation of scheduled procedures, negatively biasing the positivity rates of this group. If this bias is consistent over time, the positivity rates of this group could still be used to detect relative changes in trends. One could also circumvent this potential bias by focusing on patients admitted to the emergency room (ER) for reasons unrelated to COVID-19 (e.g., automobile accidents, lacerations, or burns). Unlike pre-procedural patients, ER patients are screened for COVID-19 onsite and lack the opportunity to modify their behaviors. Second, the demographics of pre-procedural groups may differ from the clinic population. As was observed in this study (Table 1), pre-procedural groups likely contain a greater proportion of older patients in need of corrective procedures. Weighted adjustments would be required to reconstruct a more demographically representative sample, although these correction factors may be minimal^5^. Additionally, if such distortions are consistent over time, correction factors could still allow for trend analysis and mathematical modeling of COVID-19 positivity rates. Third, the classification of patient groups in this study is based on the chief reason for testing. This method served to make the categories mutually exclusive but excluded additional factors for patients tested for multiple reasons. Although such occurrences were rare in this dataset, it is a consideration worth noting for future datasets. Fourth, some patient indications in pre-procedural groups may be correlated with COVID-19 risk. In this study, patients were tested for procedures that included colonoscopies, abdominal surgeries, joint replacements, and lacerations, which are unlikely to be correlated with COVID-19 risk. However, to further reduce this bias in future studies, pre-procedural groups may be filtered based on patient indications and history. Fifth, this study included Maui residents only, and therefore did not reflect the contributions to the positivity rate from tourism, which was expected to have increased throughout the course of this study. Increased travel was made possible by Hawaii’s Safe Travels Program, initiated on October 15, 2020, which may also have increased daily reporting due to rising tourism. However, excluding tourists from this study reduced the noise introduced by a widely diverse and fluctuating subpopulation, and allowed for a more accurate investigation of local prevalence.

### Public health implications

When COVID-19 transmission rates are sufficiently low to allow for elective procedures, pre-procedural patient datasets may be a valuable resource to reduce bias in estimates of community prevalence and improve the accuracy of critical COVID-19 metrics and trends. For example, estimating the virulence of novel variants invading communities requires COVID-19 mortality rates that are based not only on death counts, but unbiased estimates of total cases. Pre-procedural group positivity rates may also offer an additional metric to estimate the relative impact of different variants on community hospitals, as current hospital datasets often do not distinguish between patients hospitalized due to COVID-19 complications versus asymptomatic patients who are hospitalized for other medical reasons but test positive for COVID-19. Such options for reducing bias in patient sampling are essential to assess the effects of public-health interventions such as mass testing, lockdown mandates, vaccination campaigns, travel modifications, and restrictions.

## Data Availability

All data produced in the present study are available upon reasonable request to the authors.

## Acknowledgments

The authors would like to acknowledge the collaborative efforts of the Maui Medical Group staff: Cliff Alakai, Bobbi Jean Ranis, and Jessica Wallace. We thank Kristin Mills, Dr. So-One Hwang, Dr. Amanda Dolinski, and Dr. Lin H. Chen for comments. This study was commissioned by Hawaii Department of Health.

## Data and materials availability

The data used in this study were collected by the Maui Medical Group and de-identified to protect patient confidentiality.

